# Preventing the Transmission of COVID-19 in Older Adults Aged 60 Years and Above Living in Long-Term Care

**DOI:** 10.1101/2021.11.05.21265759

**Authors:** Oluwaseun Egunsola, Mark Hofmeister, Laura E. Dowsett, Tom Noseworthy, Fiona Clement

**Affiliations:** The Department Community Health Sciences, Teaching Research and Wellness Building, 3280 Hospital Drive NW Calgary Alberta T2N 4N1; O’Brien Institute for Public Health, Teaching Research and Wellness Building, 3280 Hospital Drive NW Calgary Alberta T2N 4N1

## Abstract

**Objectives:** The objective of this study was to examine the effect of measures of control and management of COVID-19, Middle East Respiratory Syndrome (MERS), and severe acute respiratory syndrome (SARS) in adults 60 years or above living in long-term care facilities. This is an update of previous work done by Rios et al.

**Methods:** A rapid review was conducted in accordance with the Rapid Review Guide for Health Policy and Systems Research. Literature search of databases MEDLINE, Cochrane library, and pre-print servers (biorxiv/medrxiv) was conducted from July 31, 2020 to October 9, 2020. EMBASE was searched from July 31, 2020 until October 18, 2020. Titles and abstracts from public archives were identified for screening using Gordon V. Cormack and Maura R. Grossman’s Continuous Active Learning® (“CAL®”) tool, which uses supervised machine learning.

**Results:** Five observational studies and one clinical practice guideline were identified. Infection prevention measures identified in this rapid review included: social distancing and isolation, personal protective equipment (PPE) use and hygiene practices, screening, training and staffing policies. The use of PPE, laboratory screening tests, sick pay to staff, self-confinement of staff within the LTCFs for 7 or more days, maintaining maximum residents’ occupancy, training and social distancing significantly reduced the prevalence of COVID-19 infection among residents and/or staff of LTCFs (p<0.05). Practices such as hiring of temporary staff, not assigning staff to care separately for infected and uninfected residents, inability to isolate sick residents and infrequent cleaning of communal areas significantly increased the prevalence of infection among residents and/or staff of LTCFs (p<0.05).

**Conclusion:** The available studies are limited to only three countries despite the global nature of the disease. The majority of these studies showed that infection control measures such as favourable staffing policies, training, screening, social distancing, isolation and use of PPE significantly improved residents and staff related outcomes. More studies exploring the effects infection prevention and control practices in long term care facilities are required.

Key Messages

- This is an update of a previous rapid review.
- Literature search of databases MEDLINE, Cochrane library, and pre-print servers (biorxiv/medrxiv) was conducted from July 31, 2020 to October 9, 2020. EMBASE was searched from July 31, 2020 until October 18, 2020.
- Five observational studies and one clinical practice guideline were identified.
- Infection prevention measures identified in this rapid review included: social distancing and isolation, PPE use and hygiene practices, screening, training and staffing policies.
- The use of PPE, laboratory screening tests, sick pay to staff, self-confinement of staff within the LTCFs for 7 or more days, maintaining maximum residents’ occupancy, training and social distancing significantly reduced the prevalence of COVID-19 infection among residents and/or staff of LTCFs (p<0.05).
- Practices such as hiring of temporary staff, not assigning staff to care separately for infected and uninfected residents, inability to isolate sick residents and infrequent cleaning of communal areas significantly increased the prevalence of infection among residents and/or staff of LTCFs (p<0.05).

## 1 Purpose

As compared to other segments of the population, older adults living in long-term care facilities have a higher risk of infection and death as a result of coronavirus 2019 (COVID-19).^1^ The overall objective of this rapid review was to examine the control and management of COVID-19, SARS, or MERS in adults 60 years or above living in long-term care facilities. This is an update of a previous work done by Rios et al.^2^ The specific research questions were:

1. What are the infection prevention and control practices for preventing or reducing the transmission of COVID-19, Middle East Respiratory Syndrome (MERS), and Severe Acute Respiratory Syndrome (SARS) in older adults aged 60 years and above living in long-term care facilities?

2. Do the infection prevention and control practices for adults aged 60 years and above living in long-term care with severe comorbidities or frailty differ as compared to those without such severe comorbidities/frailty?

3. What are the employment and remuneration policies of care providers that may have contributed to the COVID-19 outbreaks in adults aged 60 years and above living in long-term care facilities?

## 2 Methods

### 2.1 Search Strategy

A rapid review was conducted in accordance with the Rapid Review Guide for Health Policy and Systems Research.^3^ A combination of comprehensive literature searches and automated search and citation screening was used to search MEDLINE, EMBASE, Cochrane library, and pre-print servers (biorxiv/medrxiv). Grey literature was searched via international clinical trial registries (e.g., clinical trials.gov, WHO international clinical trials register), COVID-19 focused evidence gathering services (e.g., EPPI Mapper, COVID-END), as well as guideline producers/repositories (e.g., NICE guidance, ECRI).

The search for all sources for the previous review was conducted from inception up to July 31, 2020.^2^ The literature search for this update, for all sources except EMBASE, was conducted on October 9, 2020. Titles and abstracts from public archives were identified for screening using Gordon V. Cormack and Maura R. Grossman’s Continuous Active Learning® (“CAL®”) tool, which uses supervised machine learning.^4^ For archives that could be retrieved in their entirety (e.g., Medline), the entire archive was processed and searched using CAL®. For those archives that could only be accessed using keywords (e.g. clinicaltrials.gov), relevant search terms were applied (e.g., COVID-19, long-term care). The CAL® tool identifies the titles and abstracts most likely to meet specific inclusion criteria, based on the screening results that have been previously identified and reviewed. This process continues iteratively, until none of the identified articles meet the inclusion criteria. The EMBASE search was carried out from July 31, 2020 until October 18, 2020. The search strategy is available in Appendix 1. This rapid review is registered in the International Prospective Register of Systematic Reviews (CRD42020181993).

### 2.2 Study Selection

The eligibility criteria followed the PICOST framework outlined in Table 1. No other limitations were imposed. Both peer-reviewed and non-peer-reviewed papers were eligible for inclusion, as were papers written in languages other than English.

**Table 1.** Eligibility Criteria

|  |  |
| --- | --- |
| **Population** | Individuals aged $\geq 60$ years living in long-term care facilities (e.g., nursing home, long-term care hospital/facility, skilled nursing facility, convalescent home, assisted living facilities). |
| **Intervention** | Any form of infection control and prevention. Only those measures used to prevent COVID-19, MERS or SARS were included, measures related to control and prevention of other infections (e.g., vaccination for influenza, oral care to prevent bacterial pneumonia) were excluded. Additionally, interventions related to remuneration/compensation policies for long-term care facility staff, staffing models, policies on mixing of staff in long-term care facilities, and policies on staff travelling between long-term care facilities were included. |
| **Comparator** | Any of the above listed interventions listed above or no intervention |
| **Outcomes** | Lab-confirmed respiratory infection [primary outcome], symptoms, secondary transmission (e.g., other patients, healthcare workers), goal concordant care, hospitalization, intensive-care unit (ICU) admission, mortality |
| **Study designs** | Clinical practice guidelines (CPGs) and systematic reviews, using the Cochrane definition of a systematic review. Primary human studies of all designs (e.g., experimental studies, quasi-experimental studies, and observational studies excluding case series) that involved patients with COVID-19, SARS or MERS) were included. |
| **Time periods** | All periods of time and duration of follow-up were included. |

In order to meet the requested timeline, an automated approach to initial screening was used to identify the most relevant citations, the full-text of which were subsequently screened. Prior to full-text screening, calibration was conducted on five consecutive studies. Full-text screening was completed by two reviewers using Microsoft Excel. All included studies were verified by a second reviewer. A screening form based on the eligibility criteria was utilized and studies were excluded if they failed to meet the inclusion criteria as stated below.

### 2.3 Data Extraction

Items for data extraction included: title of the article, author, year of publication, country of publication, study design, inclusion criteria, total population sample, group sample sizes, number of LTCF, infection control methods and outcomes. For the clinical practice guidelines, the recommendations and level of evidence for reach recommendation was abstracted. Included studies were abstracted by a single reviewer.

### 2.4 Quality Assessment

Risk of bias appraisal was carried out by a single reviewer using the AGREE-II tool^5^ for clinical practice guidelines and the Newcastle Ottawa Scale (NOS)^6^ for cohort and case-control studies. For the NOS scale, each study was assessed across three categories: selection, comparability, and outcome. The cross-sectional studies were assessed with the Joanna Briggs Institute (JBI) checklist.^7^

### 2.5 Synthesis

The infection control interventions were classified into five broad categories: staffing policy, isolation and social distancing, personal protective equipment and hygiene, screening, and education. A narrative synthesis of the included studies was conducted.

## 3 Results

### 3.1 Study Characteristics

The search strategy yielded 457 unique citations; 376 were excluded after abstract review. Eighty-one studies proceeded to full-text review (Figure 1). Seventy-five studies were excluded for the following reasons: study design not of interest (n=35); no intervention of interest (n=33); duplicate (n=3); not retrievable (n=3); and one was an older published version of this rapid review. A total of 6 relevant studies were included in the final dataset.^8-13^

**Figure 1.**
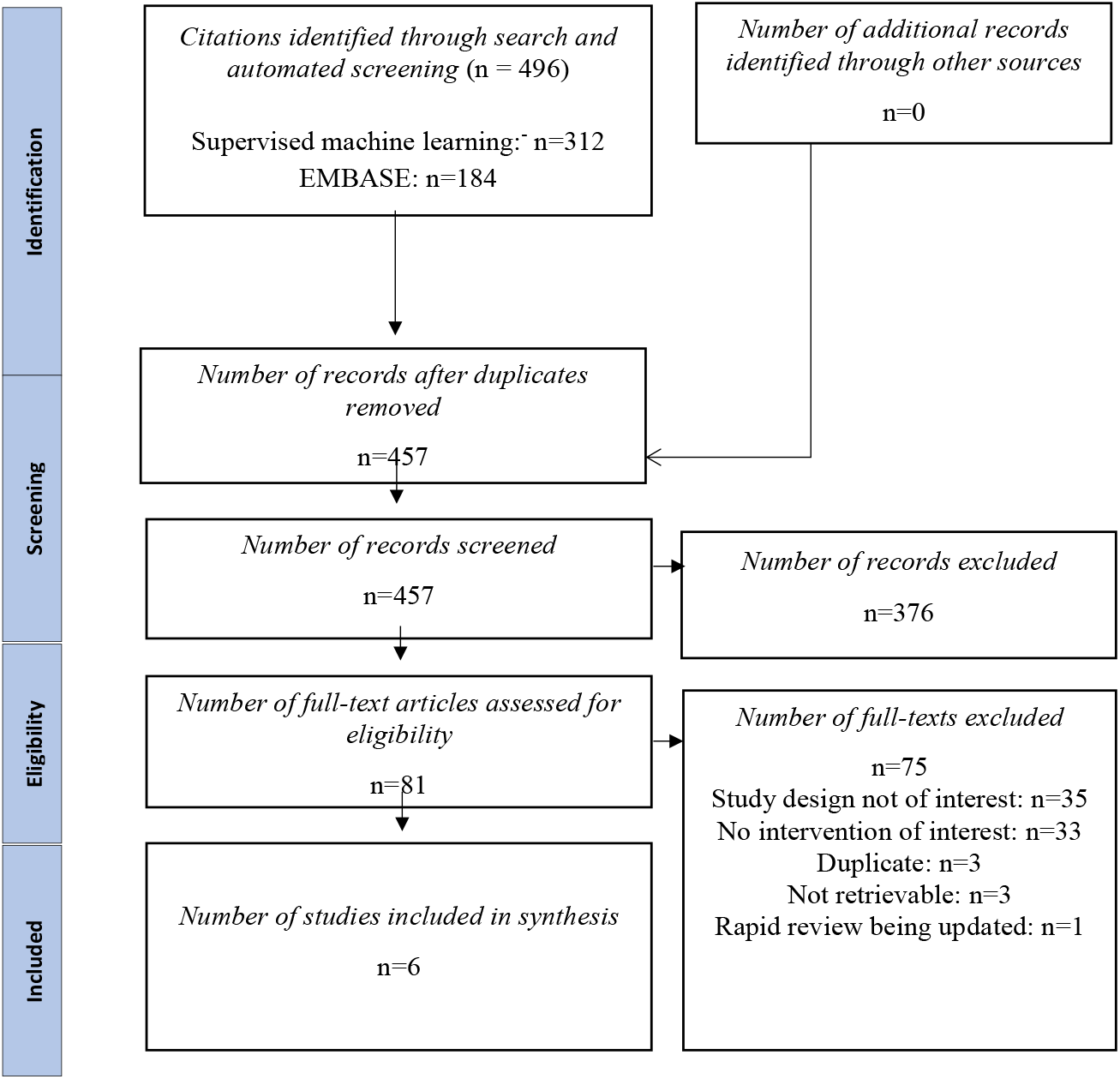
PRISMA Flowchart of Included and Excluded Studies

Two of the included studies were cohort studies,^10,11^ two were cross-sectional studies^8,9^ and one was a case-control study.^12^ One clinical practice guideline was identified.^13^ A total of 14,830 long term care facilities and 864,434 residents were involved in these studies. Two studies were conducted in France^8,10^ and the United States^11,12^, respectively and one was conducted in the United Kingdom.^9^ The only included clinical practice guideline was developed for nursing homes in Canada^13^ (Table 2).

**Table 2.** Table of Study Characteristics

| Author, Year, Country | Design | Population Sample Size | Inclusion | Infection Control Methods | Outcomes |
| --- | --- | --- | --- | --- | --- |
| Rolland, 2020<sup>8</sup><br>France | Cross-Sectional | Total Population: NR<br>Total LTCF: 124<br>Population Group 1: NR<br>Population Group 2: NR | All LTCF in Occitania Region | Staff compartmentalization<br>Resident compartmentalization<br>Use of PPE<br>Access to PPE and alcoholic sanitizers<br>Social distancing<br>Training | At least 1 case in LTCF |
| Shallcross, 2020<sup>9</sup><br>United Kingdom | Cross-Sectional | Total Population: 160,033<br>Total LTCF: 5,125<br>Population Group 1: NR<br>Population Group 2: NR | LTCF providing Care for residents with dementia or adults 65 years old or more | No temporary staffing<br>Working in single locations<br>Staff compartmentalization.<br>Payment of sick leave<br>Isolating sick residents<br>Cleaning communal areas<br>Use of PPE<br>Barrier Nursing | Prevalence of infection among residents<br>Prevalence of infection among staff<br>Proportion of LTCF with at least one case<br>Proportion of LTCF with large outbreaks |
| Belmin, 2020<sup>10</sup><br>France | Cohort | Total Population: 696,310<br>Total LTCF: 9,530<br>Population Group 1: 1,250<br>Population Group 2: 695,060 | Nursing homes in which staff voluntarily confined themselves to the facility for 7 or more days | Self-confinement by staff | At least 1 case in LTCF<br>Prevalence of infection among residents<br>Prevalence of infection among staff<br>Death rate among residents |
| Telford, 2020<sup>11</sup><br>USA | Cohort | Total Population: 5,671<br>Total LTCF: 28<br>Population Group 1: NR<br>Population Group 2: NR | Residents in LTCF | Preventive Screening | Prevalence of infection among residents<br>Prevalence of infection among staff<br>Hospitalization rate among residents |
| Telford, 2020<sup>12</sup><br>USA | Case-Control | Total Population: 2,420<br>Total LTCF: 23<br>Population Group 1: 1,150<br>Population Group 2: 1,270 | LTCF in Fulton county with at least one case of COVID-19 | Social distancing<br>Use of PPE<br>Maximum occupancy<br>Training<br>Cleaning and disinfection<br>Screening | Prevalence of infection among residents |
| Author, Year, Country | Design | Participants | Evidence Collection | Guideline Scope |  |
| Stall, 2020<sup>13</sup><br>Canada | Clinical Practice Guideline | Various authors from across Canada | 10 Provinces and 3 Territories | Guidance for reopening nursing homes for family caregivers and visitors |  |

LTCF: Long Term Care Facility, PPE: Personal Protective Equipment, NR: Not Reported

### 3.2 Quality Assessment

The two included cohort studies were truly representative of the general population and the non-exposed cohorts were from the same community as the intervention groups. In both studies, outcomes were not present at the start of the study, the cohorts were comparable, outcomes were assessed independently, with sufficient and complete follow-up. However, in one study, exposure was ascertained by self-report, thus earning no star;^11^ while exposure in the other was ascertained by structured interview.^10^ Overall, one study earned seven of eight stars^11^ while the other earned eight stars^10^ (Table 3).

**Table 3.** Quality Assessment of Non-Randomized Studies

| Cohort Studies (NOS) |  |  | Cross-Sectional Studies (JBI) |  |  | Case-Control Studies (NOS) |  |
| --- | --- | --- | --- | --- | --- | --- | --- |
| Appraisal Items | Telford<sup>11</sup> | Belmin<sup>10</sup> | Appraisal Items | Shallcross<sup>9</sup> | Rolland<sup>8</sup> |  | Telford<sup>12</sup> |
| Representative-ness of the exposed cohort | x | x | Clearly defined inclusion criteria | Yes | Yes | Case definition adequate | Yes |
| Selection of the non-exposed cohort | x | x | Subjects and setting described in detail | Yes | Yes | Representativeness of cases | Yes |
| Ascertainment of exposure |  | x | Exposure measurement valid and reliable | No | No | Selection of Controls | Yes |
| Demonstration that outcome was not present at start | x | x | Objective, standard criteria for measuring condition | Yes | Yes | Control definition | Yes |
| Comparability of cohorts (design or analysis) | x | x | Confounding factors identified | Yes | Yes | Comparability of case and controls (design or analysis) | Yes |
| Assessment of outcome | x | x | Stated strategies to deal with confounding factors | Yes | Yes | Ascertainment of exposure | No |
| Was follow-up long enough for outcomes to occur | x | x | Outcomes measured validly and reliably | Yes | Yes | Same method of ascertainment of exposure for case and control | Yes |
| Adequacy of follow up of cohorts | x | x | Appropriate statistic used | Yes | Yes | Non-response rate | No |

The case-control study was judged to be representative of the general population; the control group was from the community and had no prior history of disease. Both cases and control were comparable and the methods of ascertainment of exposure between them were comparable. There was no description of the method of ascertainment of exposure and no designation for non-response rate. Overall, this study had six of eight stars (Table 3).^12^

The two cross-sectional studies had clearly defined inclusion criteria, properly defined participants and setting, and reported objective standard for measuring COVID-19 cases. Both studies did not demonstrate the validity or reliability of the questionnaires used for exposure measurement. Both studies identified confounding factors and stated strategies to deal with them. Appropriate statistics were used in both studies^8,9^ (Table 3).

The clinical practice guideline scored four of twelve points on the scope and purpose domain, five of eleven points on the stakeholder involvement domain, no points on the rigour of development domain, two of eight points on the clarity of presentation domain, none of thirteen points on the applicability domain and one of six points on editorial independence.^13^ The full appraisal results for the included clinical practice guideline can be found in Appendix 2.

### 3.3 Narrative Synthesis of Observational Studies

#### 3.3.1 Staffing Policies

Two studies evaluated the effect of different staffing policies on COVID-19 outcomes.^8,9^ Shallcross et al. reported an almost two-fold increase in the prevalence of COVID-19 infection among residents of LTCFs employing temporary staff on most days compared with those that never employed temporary staff (p<0.001). They also found that LTCFs were about two times more likely to report a case of COVID-19 or a large outbreak of the disease if they hired temporary staff (p<0.001).^9^ Similarly, COVID-19 infection was significantly more prevalent among the staff of LTCFs that hired temporary staff than those that did not (p<0.001).^9^ However, Rolland et al. did not show any significant relationship between the proportion of LTCFs with a reported case of COVID-19 and the use of temporary staff (p=0.26).^8^ The study showed that staff compartmentalization i.e. organization of the work so that the team works in small groups in one area of the LTCF with no physical connection with the other members of the team, statistically significantly lowered the risk of COVID-19 cases in LTCFs (p=0.01).^8^ Finally, Shallcross et al. showed that providing sick pay to LTCF staff statistically significantly lowered the prevalence of infection among residents and staff (p<0.001)^9^ (Table 4, Appendix 3).

**Table 4:** Effects of Infection Control Methods and Practices on COVID-19 Outcomes in LTCFs and Residents

| Author | Practice/Infection Control Method | Outcome | Comparison | p values |
| --- | --- | --- | --- | --- |
| Rolland et al.<sup>8</sup> | *Staff compartmentalization | at least 1 case in LTCF | Yes vs No | 0.01 |
| Shallcross et al.<sup>9</sup> | Use of temporary staff | Prevalence of Infection among residents | Most days vs Never | <0.001 |
|  | *Non-cohorting of staff | Prevalence of Infection among residents | Often vs Never | <0.001 |
|  | *Non-cohorting of staff | at least 1 case in LTCF | Often vs Never | <0.001 |
|  | Sick Pay to staff | Prevalence of Infection among residents | Statutory vs None | <0.001 |
|  | Unable to isolate sick residence | Prevalence of Infection among residents | Yes vs No | <0.001 |
|  | Unable to isolate sick residence | at least 1 case in LTCF | Yes vs No | <0.001 |
|  | Cleaning communal areas | Prevalence of Infection among residents | Once vs at least twice daily | 0.003 |
|  | PPE use | Prevalence of Infection among residents | Any contact with all residents vs all the time | <0.001 |
|  | PPE use | Prevalence of Infection among residents | Direct care of all residents vs all the time | 0.007 |
|  | PPE use | Prevalence of Infection among residents | Direct care of infected residents vs all the time | <0.001 |
| Belmin et al.<sup>10</sup> | Self-confinement of staff within the facility for $\geq 7$ days | at least 1 case in LTCF | Yes vs No | <0.001 |
| | Self-confinement of staff within the facility for $\geq 7$ days | Prevalence of Infection among residents | Yes vs No | <0.001 |
| | Self-confinement of staff within the facility for $\geq 7$ days | Prevalence of death among residents | Yes vs No | <0.001 |
| Telford et al.<sup>11</sup> | Laboratory screening | Prevalence of Infection among residents | Preventive vs Responsive | <0.001 |
| Telford et al.<sup>12</sup> | Social distancing | Prevalence of infection among residents | High vs Low infection rate | 0.01 |
|  | PPE use | Prevalence of infection among residents | High vs Low infection rate | <0.001 |
|  | Maximum occupancy in LTCFs | Prevalence of infection among residents | High vs Low infection rate | 0.02 |
|  | Laminated signage about droplets | Prevalence of infection among residents | High vs Low infection rate | 0.03 |
|  | Bathroom and sink in rooms | Prevalence of infection among residents | High vs Low infection rate | 0.04 |
|  | Training and audit | Prevalence of infection among residents | High vs Low infection rate | 0.01 |

Red: Statistically significant increase in outcome; Green: Statistically significant decrease in outcome, PPE: Personal Protective Equipment; LTCF: Long-Term Care Facility; \*Non-cohorting: Staff not assigned to care separately for infected and uninfected residents; Staff compartmentalization: organization of the work so that the team works in small groups in one area of the LTCF with no physical connection with the other members of the team

#### 3.3.2 Isolation and Social Distancing

In a study of LTCFs in Fulton county, USA, social distancing and maintaining maximum occupancy limit in the facilities were statistically significantly associated with lower prevalence of COVID-19 infection (p<0.05).^12^ A French study by Belmin et al. also found significantly lower prevalence of COVID-19 infection among LTCF residents and staff, in addition to a lower prevalence of death among residents, when staff voluntarily confine themselves to the facilities for seven or more days (p<0.001).^10^ Shallcross et al. also showed that non-isolation of sick residents and non-assignment of staff to care separately for infected and uninfected residents significantly increased the risk of infections among residents and staff (p<0.001).^9^ Conversely, Rolland et al. found that compartmentalization of LTCF residents, i.e. organization of the LTCF so that they live in small groups in one area of the LTCF with no possible physical connection with the other residents; social distancing during meals and the discontinuation of group activities were not significantly associated with COVID-19 cases in the facilities ^8^ (Table 4, Appendix 3).

#### 3.3.3 Personal Protective Equipment and Personal Hygiene

When compared with using PPE all the time, Shallcross et al. found that PPE use only during contact with all or infected residents was associated with a statistically significantly lower prevalence of infection among residents and staff^9^ (Table 4). The study also showed that cleaning of communal areas less than twice daily was significantly associated with higher prevalence of COVID-19 infection among residents and staff.^9^ Similarly, Telford et al., found that nursing homes that use PPE and had bathrooms and sinks in the residents’ rooms had significantly lower infection rates (p<0.001 and p=0.04 respectively) than those that did not. They however did not find any significant relationship between cleaning and hand hygiene, and the prevalence of COVID-19 infection.^12^ Rolland et al. also did not find any significant association between PPE supply and use or the availability of hydro-alcoholic solutes and the occurrence of COVID-19 in LTCFs in France (Appendix 3).^8^

#### 3.3.4 Screening

A study by Telford et al. showed that preventive laboratory screening tests for COVID-19 in LTCFs significantly reduced the prevalence of infection among residents and staff (p<0.001). However, screening did not significantly reduce the rate of hospitalization.^11^ Another study by Telford et al. did not show any statistical association between the prevalence of COVID-19 and temperature or symptom screening (p=0.15).^12^

#### 3.3.5 Education

Telford et al. showed that LTCFs that placed signage on droplet and contact precaution in required areas reported significantly lower rates of COVID-19 infection (p=0.03). Also, those that conducted trainings and frequent audits to ensure proper mask use among staff members reported significantly fewer rates of infection (p=0.01) (Table 4).^12^

#### 3.3.6 Clinical Practice Guideline

The only clinical practice guideline included in this review was developed to guide the reopening of nursing homes to families and visitors in Canada.^13^ The guideline provided recommendations on personal protective equipment, policies for visitors, recommendations for supplies, social distancing and surveillance. Table 5 describes how this guideline compares with other available guidelines identified in the previous rapid review.^2^

**Table 5:** COVID-19 Infection control recommendations from clinical practice guidelines

| Recommendations | Stall, et al.<br>2020<sup>13</sup> | AGS, 2020 | CDC, 2020 | ECRI,<br>2020a | ECRI,<br>2020b | MOH, 2020 |
| --- | --- | --- | --- | --- | --- | --- |
| Cohorting equipment |  |  |  | X |  |  |
| Communication |  |  |  |  | X |  |
| Consulting/notifying health professionals |  | X |  | X |  | X |
| Diagnostic testing |  |  |  |  |  | X |
| Disinfecting surfaces |  |  | X | X |  | X |
| Droplet precautions |  |  |  |  |  | X |
| Education |  | X | X |  | X |  |
| Hand hygiene |  |  | X | X |  | X |
| Personal protective equipment | X |  | X | X |  | X |
| Policies for visitors | X |  | X |  |  | X |
| Policies for staff/residents |  |  |  |  |  | X |
| Provide supplies | X | X | X |  | X |  |
| Respiratory hygiene/cough etiquette |  |  | X |  | X | X |
| Social distancing/ isolation/cohorting | X | X | X | X |  |  |
| Surveillance/monitoring/evaluation | X | X | X |  | X | X |
| Compensation/sick leave policies for staff |  | X | X |  |  |  |

Grey Shaded areas adapted from Rios et al.<sup>2</sup>

## 4 Conclusion

The effect of infection prevention and control practices on COVID-19 in long-term care facilities have not been adequately explored. The available studies are limited to only three countries despite the global nature of the disease. The majority of these studies showed that infection control measures such as favourable staffing policies, training, screening, social distancing, isolation and use of PPE significantly improved residents and staff related outcomes.

## Data Availability

All datasets supporting the conclusions of this article are included within the article

## Acknowledgements

We thank Andrea Tricco and the Drug Safety and Effectiveness Network (DSEN) Methods and Applications Group for Indirect Comparisons (MAGIC) team.

## 5 Appendix

### Appendix 1

#### EMBASE search strategy

1. exp coronaviridae/ or exp Coronaviridae infection/ or exp Coronavirus infection/ or SARS coronavirus/
2. ((wuhan or hubei or huanan) and (severe acute respiratory or pneumonia^*^ or virus^*^) and outbreak^*^).mp.
3. (coronavir^*^ or “corona virus^*^” or “coronavirus pneumonia” or betacoronavir^*^ or COVID or COVID-19).mp.
4. (“nCoV” or “cov 2” or cov2 or 2019ncov or 2019-nCoV or “2019 ncov” or “2019-ncov” or “2019 novel cov” or “2019 ncov disease^*^” or “2019 novel coronavirus^*^”).mp.
5. (“severe acute respiratory syndrome coronavirus^*^” or “wuhan virus^*^” or “sars cov 2 mers” or “middle east respiratory syndrome^*^” or “Severe Acute Respiratory” or SARS or SARS-CoV or SARScov2 or MERS-CoV).mp.
6. or/1-5
7. exp communicable disease control/ or exp “prevention and control”/
8. contact examination/
9. exp protective equipment/ or exp surgical attire/
10. exp hygiene/ or exp hand washing/
11. patient isolation/ or contact examination/
12. instrument sterilization/ or exp disinfection/ or decontamination/
13. bleaching agent/
14. (“infection control” or “virus control” or “disease control” or prevent^*^ or handwash^*^ or “hand wash^*^” or quarant^*^ or isolat^*^ or steril^*^ or disinfect^*^ or fumigat^*^ or decontaminat^*^ or resanitiz^*^ or resanitis^*^ or desaniti^*^ or contaminat^*^ or antisept^*^ or biocid^*^ or steriliz^*^ or sanitize^*^ or bleach^*^ or hypochlor^*^ or ozon^*^ or ultraviolet or UV or “contract tracing” or “disease notification”).mp.
15. (“protective equipment” or “protective cloth^*^” or “protective product^*^” or “protective gear” or PPE or PPEs or mask^*^ or facemask^*^ or half-mask^*^ or facepiece^*^ or n95^*^ or n99^*^ or shield^*^ or faceshield^*^ or “Particulate filter^*^” or “gas filter^*^” or glov^*^ or gown or gowns or “space suits” or “respiratory protect^*^” or visor or “eye protect^*^ “or “eye spectacle^*^ “or “hand protect^*^ “or “hand wash^*^” or “handwash^*^” or google or goggles or “head cover^*^ “or “shoe cover^*^” or respirator^*^ or ventilator^*^).mp.
16. (restrict^*^ adj3 (resident^*^ or patient^*^ or visit^*^ or family or travel^*^ or staff or provider^*^ or employee^*^)).mp.
17. ((respiratory or cough or hand) adj2 (hygiene or etiquette)).mp.
18. exp ventilator/
19. or/7-18
20. 6 and 19
21. nursing home/ or home for the aged/ or assisted living facility/
22. ((elder^*^ or senior or nursing or aged or “old age” or “old people” or “old person^*^” or “long-term care” or “LTC” or “long term care”) adj2 (home or homes or hous^*^ or residenc^*^ or facilit^*^ or hospital^*^)).mp.
23. (“convalescence hom^*^” or “convalescence hospital^*^” or “extended care facility^*^” or “charitable hom^*^” or “home based health care facilit^*^”).mp.
24. exp long term care/
25. or/21-24
26. 20 and 25

### Appendix 2

AGREE checklist for Stall et al.^13^

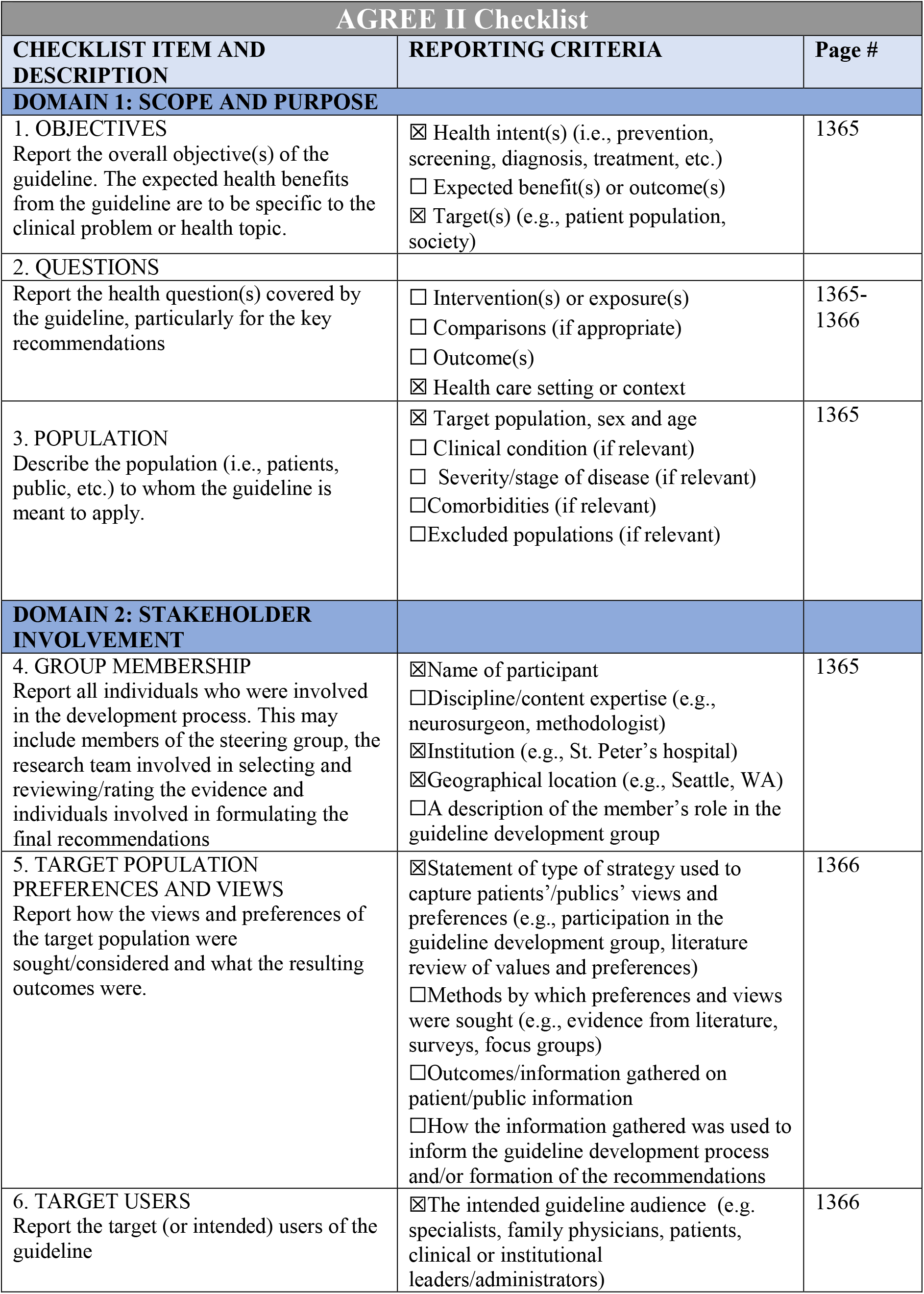
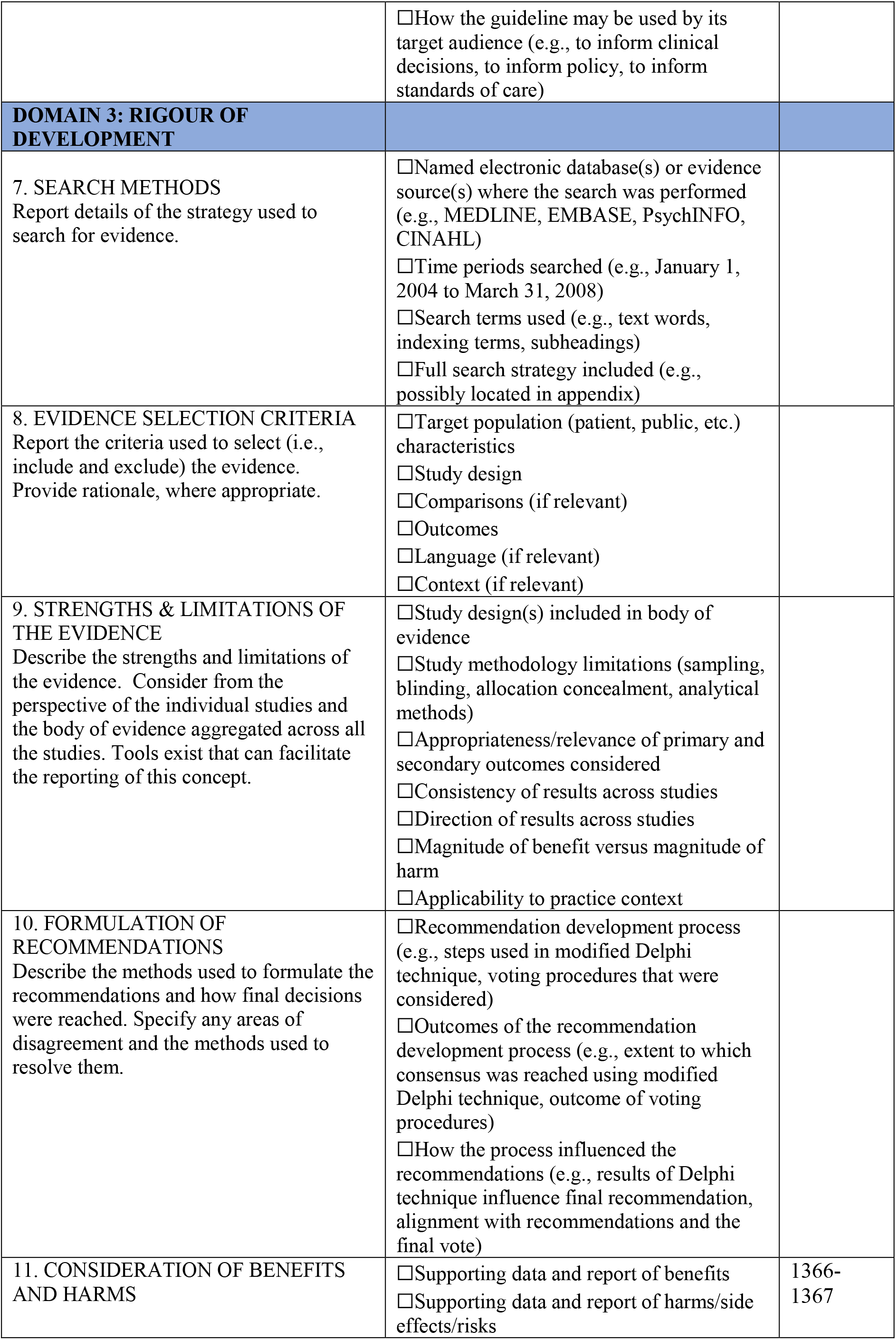
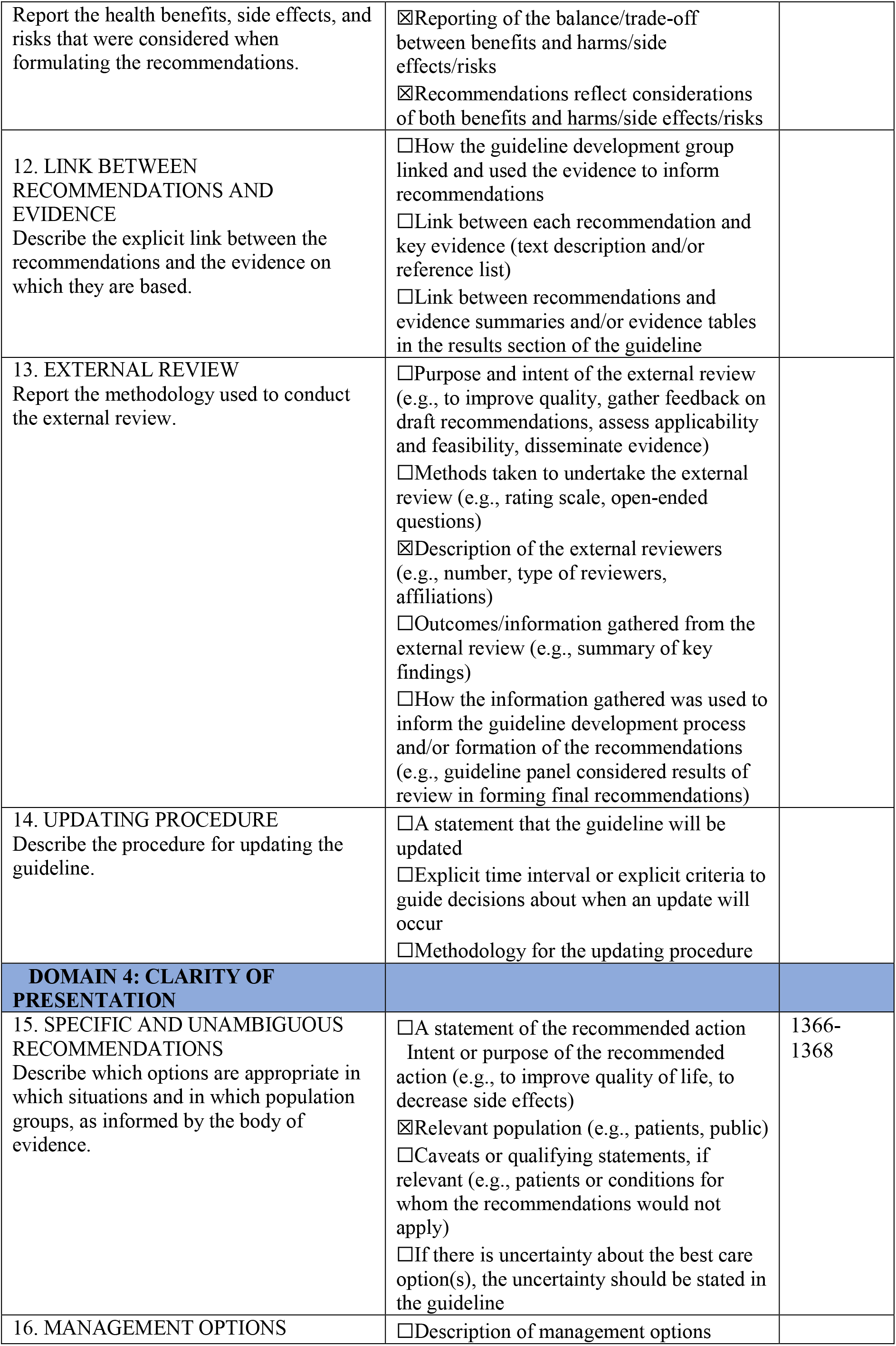
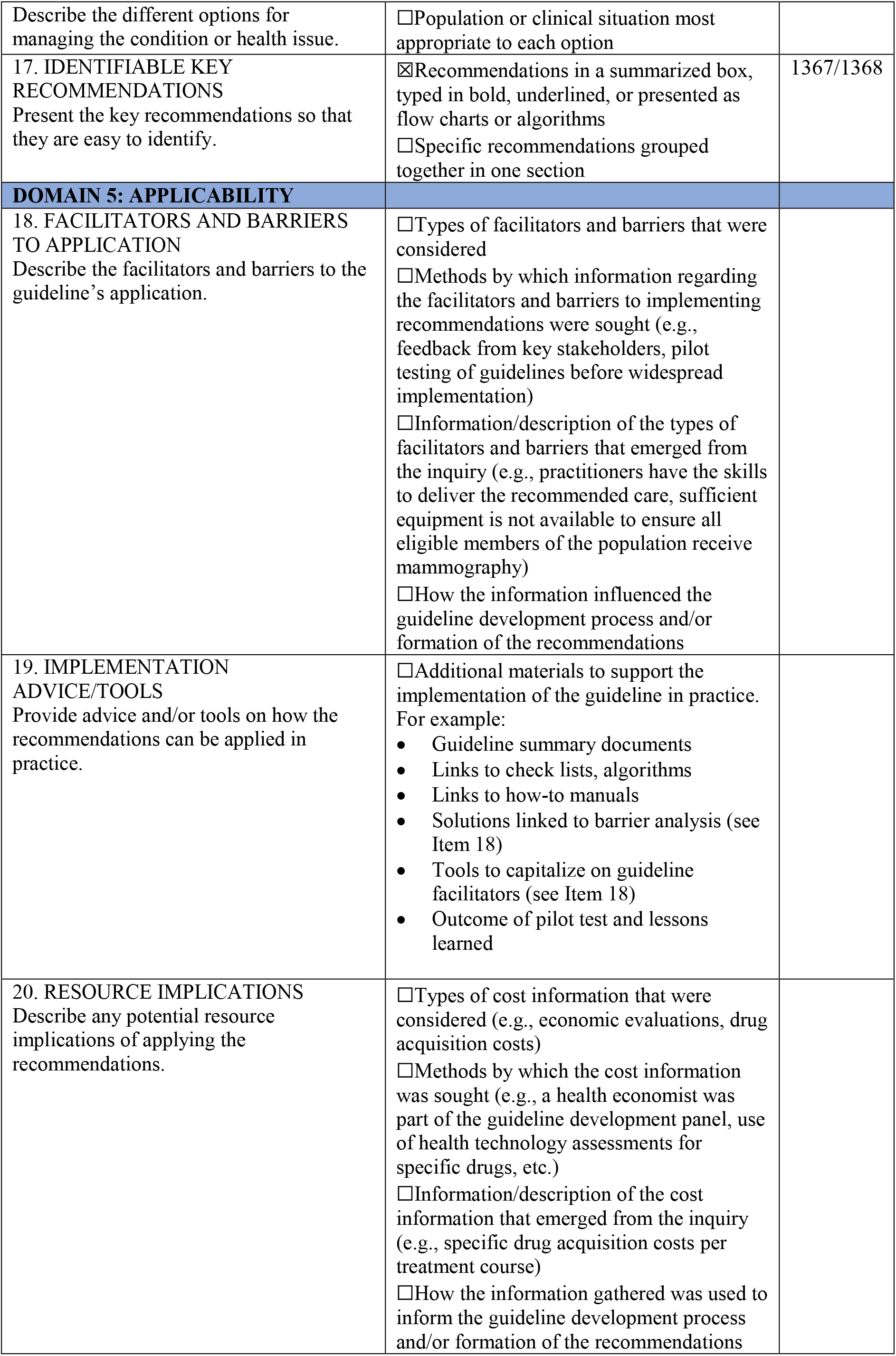
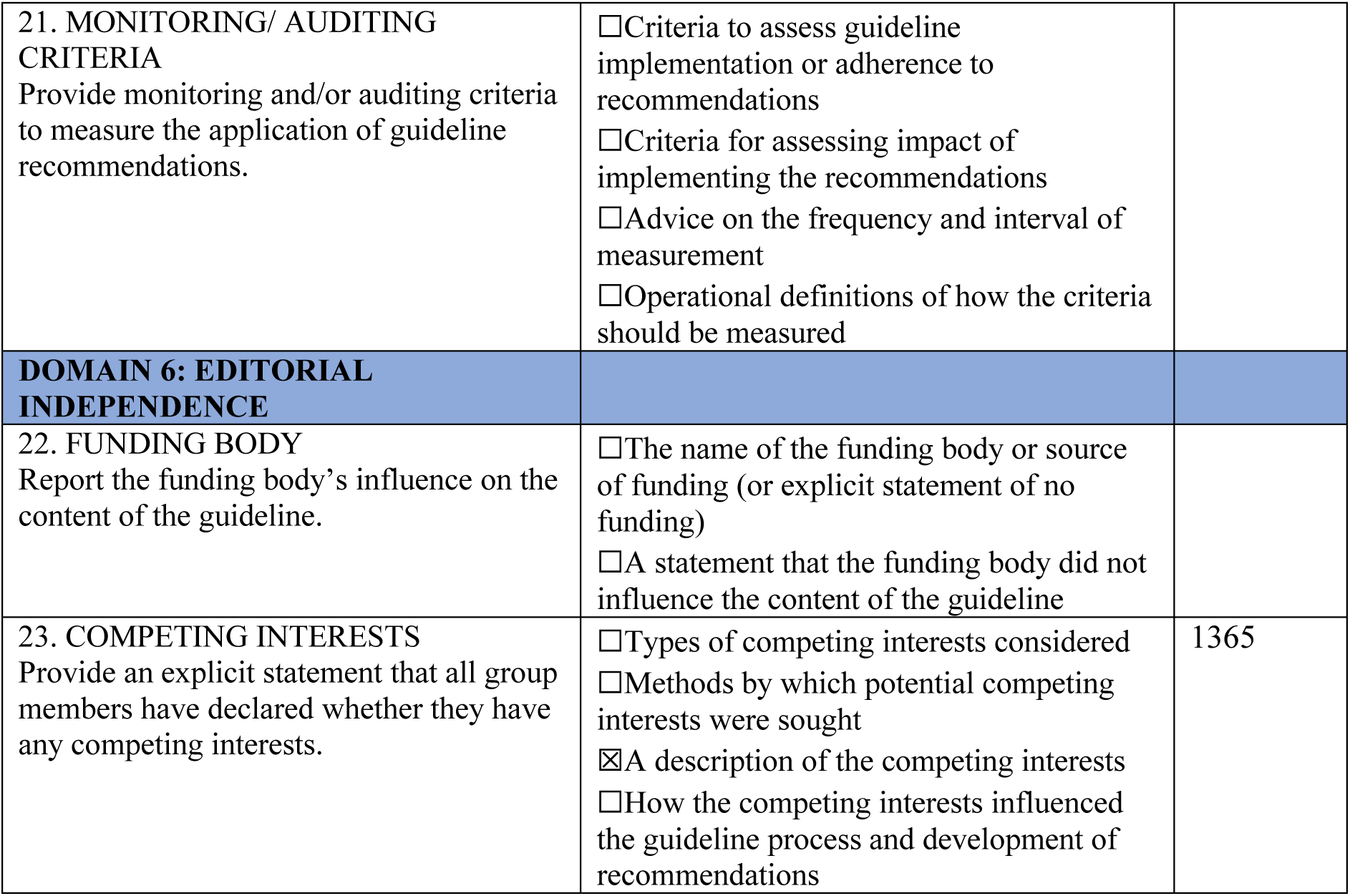

### Appendix 3

Table of study characteristics with quantitative results

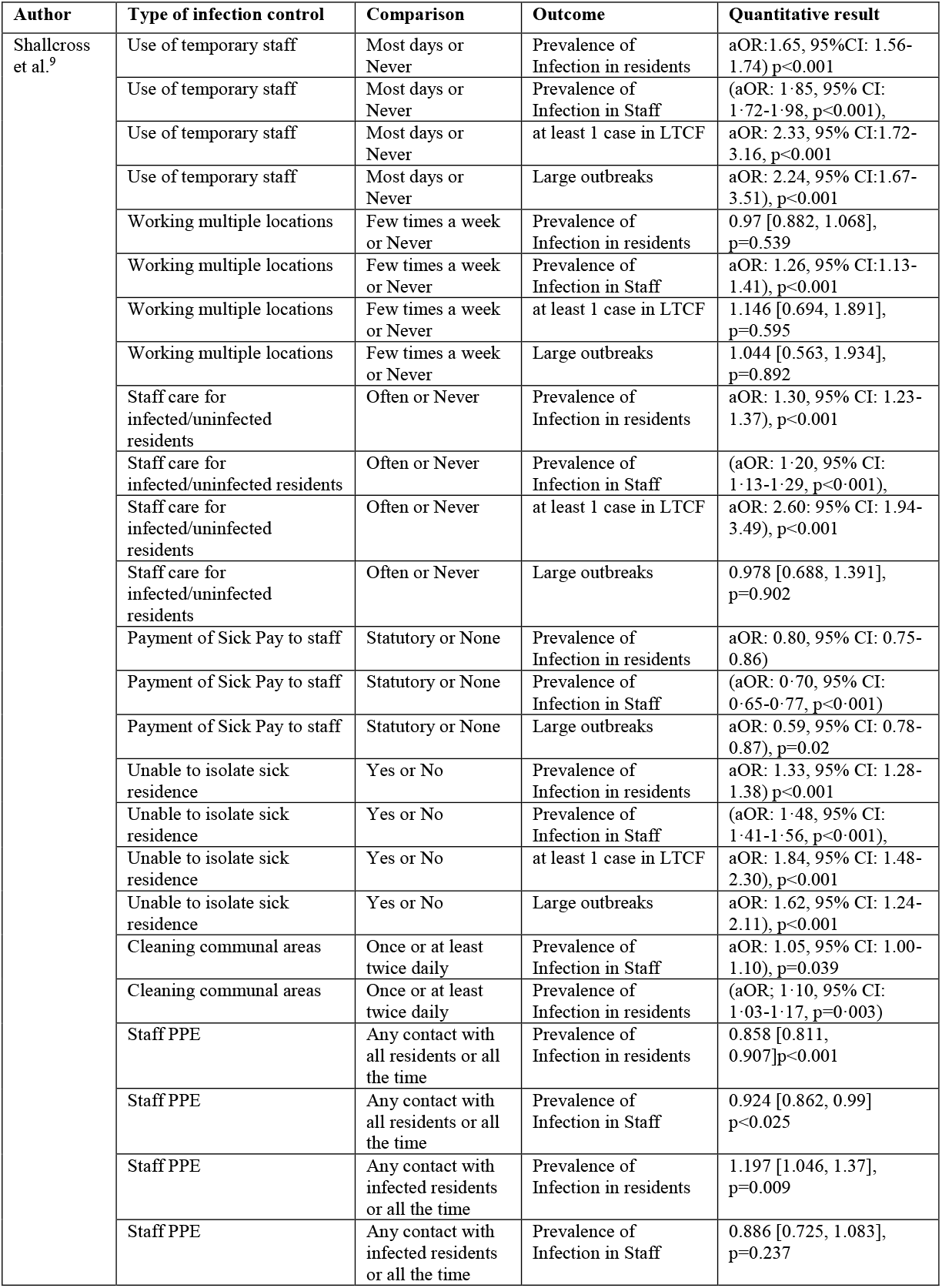
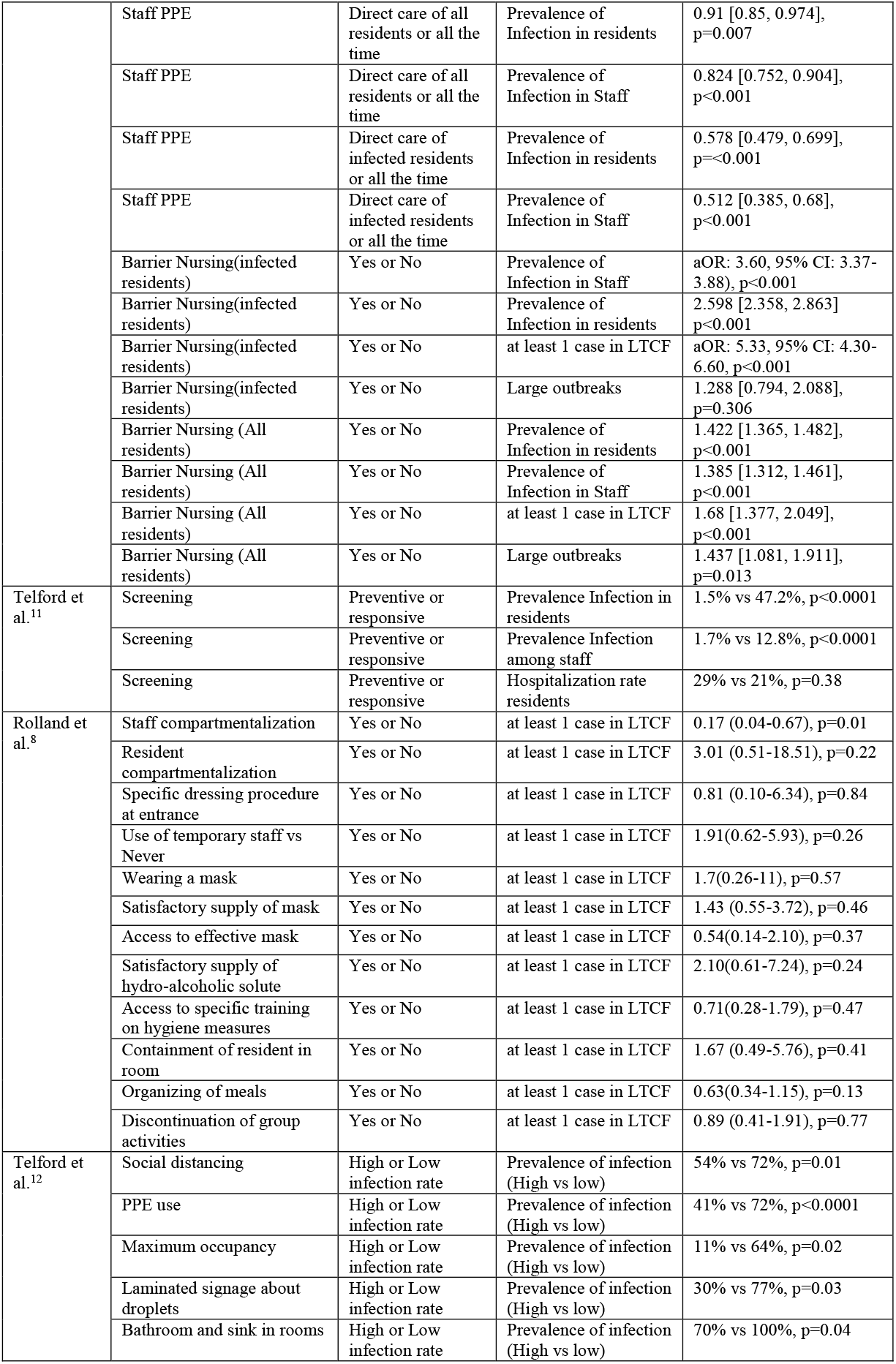
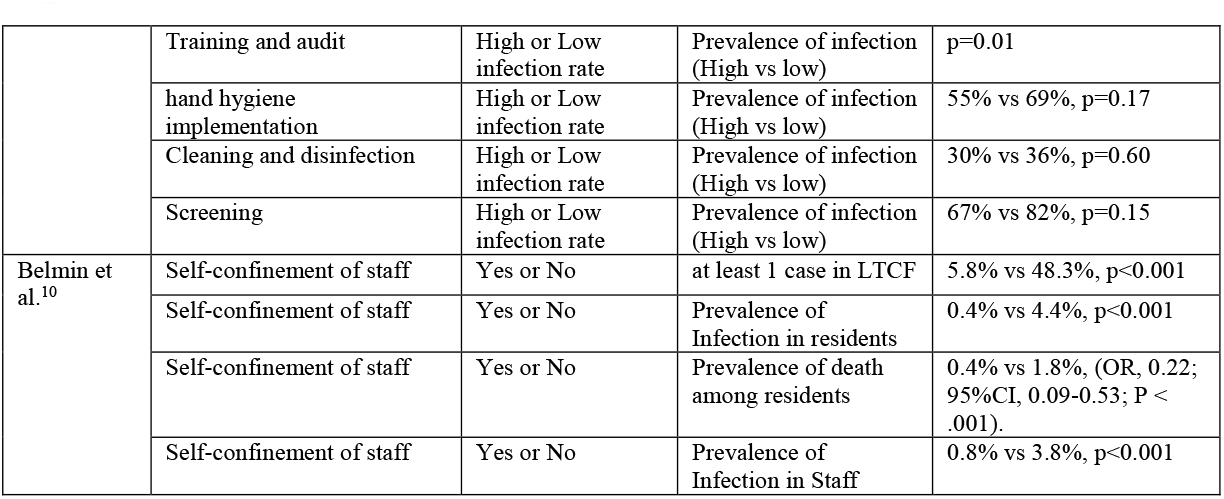

Appendix 3 Table of study characteristics with quantitative results
| Author | Type of infection control | Comparison | Outcome | Quantitative result |
| --- | --- | --- | --- | --- |
| Shallcross et al.<sup>9</sup> | Use of temporary staff | Most days or Never | Prevalence of Infection in residents | aOR: 1.65, 95% CI: 1.56-1.74) p<0.001 |
|  | Use of temporary staff | Most days or Never | Prevalence of Infection in Staff | (aOR: 1.85, 95% CI: 1.72-1.98, p<0.001), |
|  | Use of temporary staff | Most days or Never | at least 1 case in LTCF | aOR: 2.33, 95% CI: 1.72-3.16, p<0.001 |
|  | Use of temporary staff | Most days or Never | Large outbreaks | aOR: 2.24, 95% CI: 1.67-3.51), p<0.001 |
|  | Working multiple locations | Few times a week or Never | Prevalence of Infection in residents | 0.97 [0.882, 1.068], p=0.539 |
|  | Working multiple locations | Few times a week or Never | Prevalence of Infection in Staff | aOR: 1.26, 95% CI: 1.13-1.41), p<0.001 |
|  | Working multiple locations | Few times a week or Never | at least 1 case in LTCF | 1.146 [0.694, 1.891], p=0.595 |
|  | Working multiple locations | Few times a week or Never | Large outbreaks | 1.044 [0.563, 1.934], p=0.892 |
|  | Staff care for infected/uninfected residents | Often or Never | Prevalence of Infection in residents | aOR: 1.30, 95% CI: 1.23-1.37), p<0.001 |
|  | Staff care for infected/uninfected residents | Often or Never | Prevalence of Infection in Staff | (aOR: 1.20, 95% CI: 1.13-1.29, p<0.001), |
|  | Staff care for infected/uninfected residents | Often or Never | at least 1 case in LTCF | aOR: 2.60: 95% CI: 1.94-3.49), p<0.001 |
|  | Staff care for infected/uninfected residents | Often or Never | Large outbreaks | 0.978 [0.688, 1.391], p=0.902 |
|  | Payment of Sick Pay to staff | Statutory or None | Prevalence of Infection in residents | aOR: 0.80, 95% CI: 0.75-0.86) |
|  | Payment of Sick Pay to staff | Statutory or None | Prevalence of Infection in Staff | (aOR: 0.70, 95% CI: 0.65-0.77, p<0.001) |
|  | Payment of Sick Pay to staff | Statutory or None | Large outbreaks | aOR: 0.59, 95% CI: 0.78-0.87), p=0.02 |
|  | Unable to isolate sick residence | Yes or No | Prevalence of Infection in residents | aOR: 1.33, 95% CI: 1.28-1.38) p<0.001 |
|  | Unable to isolate sick residence | Yes or No | Prevalence of Infection in Staff | (aOR: 1.48, 95% CI: 1.41-1.56, p<0.001), |
|  | Unable to isolate sick residence | Yes or No | at least 1 case in LTCF | aOR: 1.84, 95% CI: 1.48-2.30), p<0.001 |
|  | Unable to isolate sick residence | Yes or No | Large outbreaks | aOR: 1.62, 95% CI: 1.24-2.11), p<0.001 |
|  | Cleaning communal areas | Once or at least twice daily | Prevalence of Infection in Staff | aOR: 1.05, 95% CI: 1.00-1.10), p=0.039 |
|  | Cleaning communal areas | Once or at least twice daily | Prevalence of Infection in residents | (aOR: 1.10, 95% CI: 1.03-1.17, p=0.003) |
|  | Staff PPE | Any contact with all residents or all the time | Prevalence of Infection in residents | 0.858 [0.811, 0.907]p<0.001 |
|  | Staff PPE | Any contact with all residents or all the time | Prevalence of Infection in Staff | 0.924 [0.862, 0.99] p<0.025 |
|  | Staff PPE | Any contact with infected residents or all the time | Prevalence of Infection in residents | 1.197 [1.046, 1.37], p=0.009 |
|  | Staff PPE | Any contact with infected residents or all the time | Prevalence of Infection in Staff | 0.886 [0.725, 1.083], p=0.237 |
|  | Staff PPE | Direct care of all residents or all the time | Prevalence of Infection in residents | 0.91 [0.85, 0.974], p=0.007 |
|  | Staff PPE | Direct care of all residents or all the time | Prevalence of Infection in Staff | 0.824 [0.752, 0.904], p<0.001 |
|  | Staff PPE | Direct care of infected residents or all the time | Prevalence of Infection in residents | 0.578 [0.479, 0.699], p<0.001 |
|  | Staff PPE | Direct care of infected residents or all the time | Prevalence of Infection in Staff | 0.512 [0.385, 0.68], p<0.001 |
|  | Barrier Nursing(infected residents) | Yes or No | Prevalence of Infection in Staff | aOR: 3.60, 95% CI: 3.37-3.88), p<0.001 |
|  | Barrier Nursing(infected residents) | Yes or No | Prevalence of Infection in residents | 2.598 [2.358, 2.863] p<0.001 |
|  | Barrier Nursing(infected residents) | Yes or No | at least 1 case in LTCF | aOR: 5.33, 95% CI: 4.30-6.60, p<0.001 |
|  | Barrier Nursing(infected residents) | Yes or No | Large outbreaks | 1.288 [0.794, 2.088], p=0.306 |
|  | Barrier Nursing (All residents) | Yes or No | Prevalence of Infection in residents | 1.422 [1.365, 1.482], p<0.001 |
|  | Barrier Nursing (All residents) | Yes or No | Prevalence of Infection in Staff | 1.385 [1.312, 1.461], p<0.001 |
|  | Barrier Nursing (All residents) | Yes or No | at least 1 case in LTCF | 1.68 [1.377, 2.049], p<0.001 |
|  | Barrier Nursing (All residents) | Yes or No | Large outbreaks | 1.437 [1.081, 1.911], p=0.013 |
| Telford et al.<sup>11</sup> | Screening | Preventive or responsive | Prevalence Infection in residents | 1.5% vs 47.2%, p<0.0001 |
|  | Screening | Preventive or responsive | Prevalence Infection among staff | 1.7% vs 12.8%, p<0.0001 |
|  | Screening | Preventive or responsive | Hospitalization rate residents | 29% vs 21%, p=0.38 |
| Rolland et al.<sup>8</sup> | Staff compartmentalization | Yes or No | at least 1 case in LTCF | 0.17 (0.04-0.67), p=0.01 |
|  | Resident compartmentalization | Yes or No | at least 1 case in LTCF | 3.01 (0.51-18.51), p=0.22 |
|  | Specific dressing procedure at entrance | Yes or No | at least 1 case in LTCF | 0.81 (0.10-6.34), p=0.84 |
|  | Use of temporary staff vs Never | Yes or No | at least 1 case in LTCF | 1.91(0.62-5.93), p=0.26 |
|  | Wearing a mask | Yes or No | at least 1 case in LTCF | 1.7(0.26-11), p=0.57 |
|  | Satisfactory supply of mask | Yes or No | at least 1 case in LTCF | 1.43 (0.55-3.72), p=0.46 |
|  | Access to effective mask | Yes or No | at least 1 case in LTCF | 0.54(0.14-2.10), p=0.37 |
|  | Satisfactory supply of hydro-alcoholic solute | Yes or No | at least 1 case in LTCF | 2.10(0.61-7.24), p=0.24 |
|  | Access to specific training on hygiene measures | Yes or No | at least 1 case in LTCF | 0.71(0.28-1.79), p=0.47 |
|  | Containment of resident in room | Yes or No | at least 1 case in LTCF | 1.67 (0.49-5.76), p=0.41 |
|  | Organizing of meals | Yes or No | at least 1 case in LTCF | 0.63(0.34-1.15), p=0.13 |
|  | Discontinuation of group activities | Yes or No | at least 1 case in LTCF | 0.89 (0.41-1.91), p=0.77 |
| Telford et al.<sup>12</sup> | Social distancing | High or Low infection rate | Prevalence of infection (High vs low) | 54% vs 72%, p=0.01 |
|  | PPE use | High or Low infection rate | Prevalence of infection (High vs low) | 41% vs 72%, p<0.0001 |
|  | Maximum occupancy | High or Low infection rate | Prevalence of infection (High vs low) | 11% vs 64%, p=0.02 |
|  | Laminated signage about droplets | High or Low infection rate | Prevalence of infection (High vs low) | 30% vs 77%, p=0.03 |
|  | Bathroom and sink in rooms | High or Low infection rate | Prevalence of infection (High vs low) | 70% vs 100%, p=0.04 |
|  | Training and audit | High or Low infection rate | Prevalence of infection (High vs low) | p=0.01 |
|  | hand hygiene implementation | High or Low infection rate | Prevalence of infection (High vs low) | 55% vs 69%, p=0.17 |
|  | Cleaning and disinfection | High or Low infection rate | Prevalence of infection (High vs low) | 30% vs 36%, p=0.60 |
|  | Screening | High or Low infection rate | Prevalence of infection (High vs low) | 67% vs 82%, p=0.15 |
| Belmin et al.<sup>10</sup> | Self-confinement of staff | Yes or No | at least 1 case in LTCF | 5.8% vs 48.3%, p<0.001 |
|  | Self-confinement of staff | Yes or No | Prevalence of Infection in residents | 0.4% vs 4.4%, p<0.001 |
|  | Self-confinement of staff | Yes or No | Prevalence of death among residents | 0.4% vs 1.8%, (OR, 0.22; 95%CI, 0.09-0.53; P < .001). |
|  | Self-confinement of staff | Yes or No | Prevalence of Infection in Staff | 0.8% vs 3.8%, p<0.001 |

## References

1. Fisman DN, Bogoch I, Lapointe-Shaw L, McCready J, Tuite AR. Risk factors associated with mortality among residents with coronavirus disease 2019 (COVID-19) in long-term care facilities in Ontario, Canada. JAMA network open. 2020;3(7):e2015957–e2015957.

2. Rios P, Radhakrishnan A, Williams C, et al. Preventing the transmission of COVID-19 and other coronaviruses in older adults aged 60 years and above living in long-term care: a rapid review. Systematic Reviews. 2020;9(1):1–8.

3. Tricco AC, Langlois E, Straus SE, Organization WH. Rapid reviews to strengthen health policy and systems: a practical guide. World Health Organization; 2017.

4. Cormack GV, Grossman MR. Technology-Assisted Review in Empirical Medicine: Waterloo Participation in CLEF eHealth 2017. Paper presented at: CLEF (Working Notes) 2017.

5. AGREE. AGREE-II Reporting Checklist. https://www.agreetrust.org/resource-centre/agree-reporting-checklist/. Published 2016. Accessed 26 October 2020.

6. Wells GA, Shea B, O’Connell D, et al. The Newcastle-Ottawa Scale (NOS) for assessing the quality of nonrandomised studies in meta-analyses. In: Oxford; 2000.

7. Joanna Briggs Institute. Checklist for analytical cross sectional studies. 2020. https://joannabriggs.org/sites/default/files/2020-08/Checklist_for_Analytical_Cross_Sectional_Studies.pdf. Accessed 26 October 2020.

8. Rolland Y, Lacoste M-H, De Mauleon A, et al. Guidance for the Prevention of the COVID-19 Epidemic in Long-Term Care Facilities: A Short-Term Prospective Study. The journal of nutrition, health & aging. 2020:1–5.

9. Shallcross L, Burke D, Abbott O, et al. Risk factors associated with SARS-CoV-2 infection and outbreaks in Long Term Care Facilities in England: a national survey. medRxiv. 2020.

10. Belmin J, Um-Din N, Donadio C, et al. Coronavirus Disease 2019 Outcomes in French Nursing Homes That Implemented Staff Confinement With Residents. JAMA network open. 2020;3(8):e2017533–e2017533.

11. Telford CT, Onwubiko U, Holland D, et al. Mass Screening for SARS-CoV-2 Infection among Residents and Staff in Twenty-eight Long-term Care Facilities in Fulton County, Georgia. medRxiv. 2020.

12. Telford CT, Bystrom C, Fox T, et al. Assessment of infection prevention and control protocols, procedures, and implementation in response to the COVID-19 pandemic in twenty-three long-term care facilities in Fulton County, Georgia. medRxiv. 2020.

13. Stall NM, Johnstone J, McGeer AJ, Dhuper M, Dunning J, Sinha SK. Finding the Right Balance: An Evidence-Informed Guidance Document to Support the Re-Opening of Canadian Nursing Homes to Family Caregivers and Visitors during the Coronavirus Disease 2019 Pandemic. Journal of the American Medical Directors Association. 2020;21(10):1365-1370. e1367.

